# Perceptions and experiences of rare diseases among General Practitioners: an exploratory study

**DOI:** 10.1101/2021.09.07.21263025

**Authors:** Julie McMullan, Ashleen L. Crowe, Taylor McClenaghan, Helen McAneney, Amy Jayne McKnight

## Abstract

**Background:** General Practitioners (GPs) play an important role in healthcare delivery for people living with a rare disease and their families. Managing long-term multisystem diseases that often have no cure and sub-optimal treatment options can be extremely challenging.

**Aim:** To gain an understanding of GP’s perception and experience of rare diseases.

**Design and Setting:** This is an exploratory study with GPs from Northern Ireland.

**Method:** An online survey was hosted within SmartSurvey, available for 6 weeks in early 2019, which queried: GPs career to date, interactions with rare disease patients, challenges faced, the exchange of information with patients, priorities for the future, support for such patients and finally training.

**Results:** Sixty-six GPs engaged with the survey with 15 completing the survey. Many frustrations were mentioned by GPs when caring for such patients including finding a diagnosis, lack of specialist services and lack of awareness. Seventy-nine percent of GPs responding did not feel adequately trained to care for patients with a rare disease, while 93% were interested in undertaking training to enable them to improve their care for rare disease patients.

**Conclusion:** More effective and convenient ‘rare disease’ focused training programs would help GPs manage patients with rare diseases. Enabling efficient, effective communication between GPs and individual medical specialists is of paramount importance to give GPs access to information on how to effectively manage patients living with a rare disease. Awareness should be raised for effective support mechanisms such as charities and support groups for rare disease patients.

**How this fits in:** Many GPs have reported feeling overwhelmed when caring for rare disease patients. Their lack of knowledge forces them to turn to the internet but finding reliable information is often difficult. This study acknowledges the difficult task GPs face when caring for this population and highlights the need for training development, improved communication, and better awareness.

## Introduction

A rare disease is defined by the European Medicines Agency as a ‘life-threatening or chronically debilitating’ condition which affects no more than 5 per 10000 people in the EU (1). In recent years rare diseases have been acknowledged as a major public health concern (2). There are approximately 8000 known rare diseases, affecting around 25 to 30 million people in Europe (3);many of these diseases are unknown to the majority of healthcare professionals (HCPs) (3). Managing rare diseases in clinical practice is difficult due to their diversity, their complexity, and lack of dedicated rare disease training (4).

Due to the low prevalence and lack of expertise for many conditions, patients living with a rare disease are often compelled to become knowledgeable about their own disease. They become ‘expert patients’ alongside their carer(s) as they seek an empowering and collaborative approach with their clinicians (5-8). This is a shift from the traditional patient-doctor relationship with each participant revising their role and expectations (8). An accessible, proactive General Practitioner (GP) relationship may help reduce many of the negative aspects of living with a rare disease (9) as well as lowering medical costs and minimising misuse of medical service (e.g. fewer diagnostic tests), by adopting a patient centred approach to care (10, 11).

Previous research indicates that those with a rare disease experience difficulties finding relatability, empathy, relevant knowledge, adequate communication, and understanding from the HCPs they encounter (7, 12-14). GPs play an important role in healthcare delivery for people living with a rare disease and their families by increasing familiarity with the disease and the impact it is likely to have on the patient and their support network. (2, 5, 15). GPs are often the first point of contact for those presenting with symptoms and when diagnosed, rare disease patients will attend a GP between visits to specialist services. GPs may take an active role in developing and implementing appropriate care plans and case coordination for rare disease patients (16).

The *Northern Ireland Rare Disease Implementation Plan (2015)* (17) recognised that GPs need further training to facilitate effective diagnosis and intervention to treat people with rare diseases, and that protocols for those who have no diagnosis or are at potential risk of having a rare disease should be put in place. The *UK strategy for Rare Diseases* (18) emphasised the need for improvement of the coordination of care, diagnosis, treatment and patient empowerment.

The exploratory approach undertaken in this study explored GPs perceptions and experiences of rare diseases within a GP population from Northern Ireland.

## Methods

An exploration was conducted of the experience of GP’s interactions with patients living with a rare disease(s) across Northern Ireland (NI) *via* an online survey. A primarily qualitative approach was chosen (19) for this study as the aim was to gain insight into the perceptions and experiences of GPs regarding rare diseases. Qualitative research aims to understand behaviour and beliefs, identify processes and to understand the context of people’s experiences (19). This study adopts an interpretivism approach and has a strong commitment to constructionist ontology (20, 21). This paper has been written in accordance with the guidance for reporting qualitative research as outlined by O’Brien *et al*. (2014) (22).

Ethical approval was provided by the Faculty of Medicine, Life and Health Sciences, Queen’s University of Belfast (QUB), (reference 18.47v3). Results were collated anonymously and used to evaluate current knowledge of GPs and identify where additional resources are needed to better support them.

An online survey was hosted within SmartSurvey (www.smartsurvey.co.uk) (23) [from mid January 2019 for 6 weeks] with two sections and 26 questions (Supplementary file 1). The survey was constructed using an iterative approach with input from the Northern Ireland Rare Disease Partnership (NIRDP), the national charity for rare diseases in NI (24) and was uploaded to the Smart Survey platform (23). The survey queried: GPs career to date, interactions with rare disease patients, challenges faced, the exchange of information with patients, priorities for the future, support for such patients and finally training. The survey was comprised of closed and open-ended questions and respondents had the option to save and complete the questions at a later date.

A weblink to the survey and participant information leaflet were advertised *via* a variety of online platforms including the NIRDP website (www.nirdp.org.uk) (24). The survey link was posted on Twitter on a recruitment notice. It was also submitted for inclusion in a NI Royal College of General Practitioners newsletter, and QUBComms (www.qub.ac.uk/sites/StaffGateway/RoundUp/) (25) newsfeed. To be eligible to participate, potential respondents had to be a GP currently working in NI.

SmartSurvey (23) was used to generate tables and charts to add demographic and statistical information to the study. Worditout.com (26) was used to create a word cloud (Supplementary figure 1) incorporating the views taken from question 12 of the survey. It illustrates the biggest challenges GPs face when caring for rare disease patients.

## Results

### Respondent characteristics

Sixty-six GPs participated in the survey with 15 proceeding through questions in the survey and pressing the ‘submit’ button. Given the time constraints and pressures GPs find themselves under, this was considered a reasonable number. The demographics of those who responded fully are displayed in Table 1. 73.3% of respondents were female, over half were from Northern Ireland (53.3%) and 46.2% had been working as a GP for 0-10 years at the time of the survey.

**Table 1.**
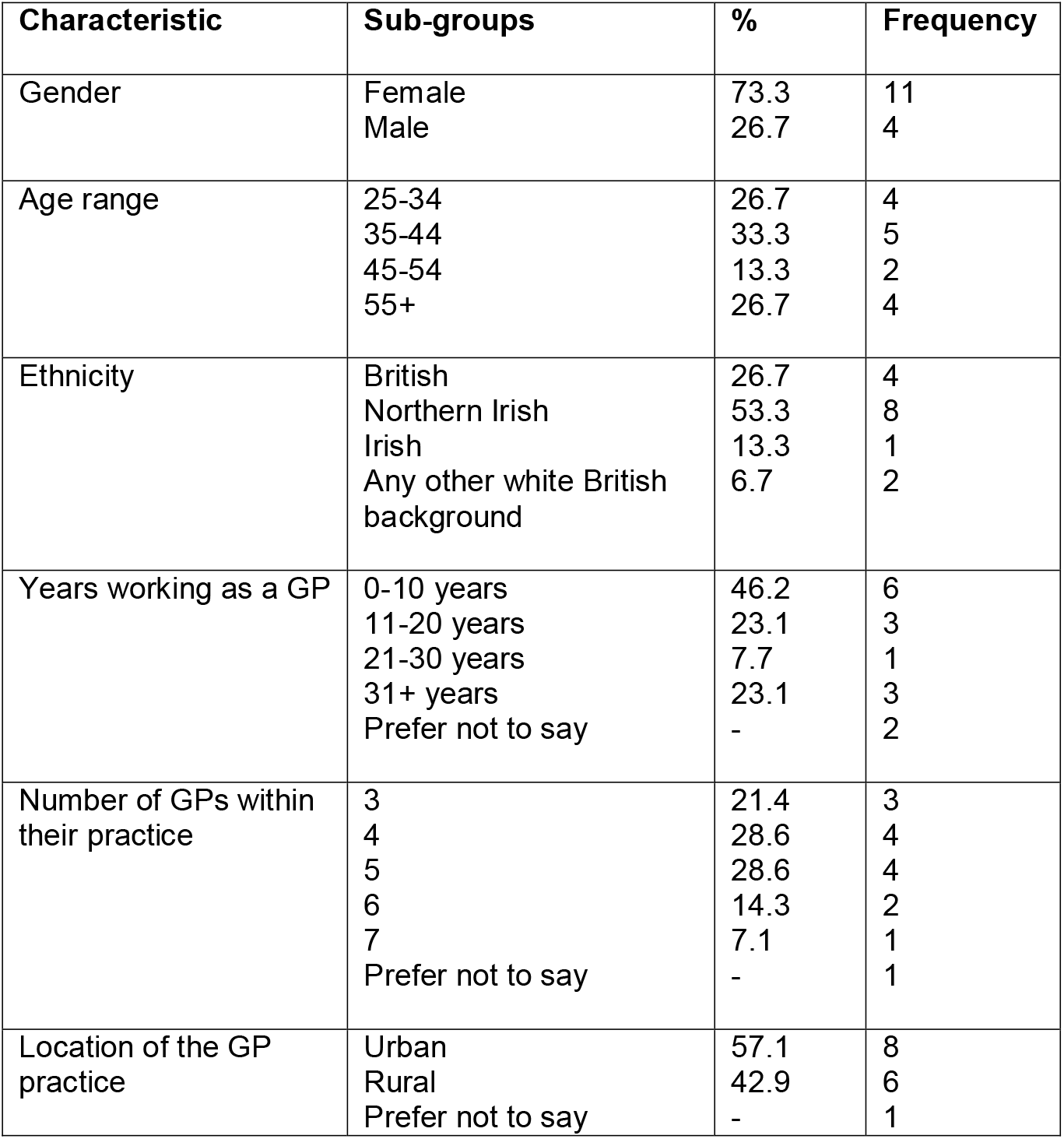
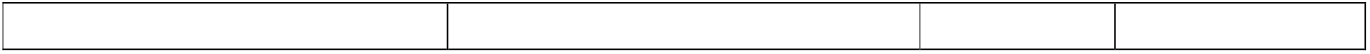
Demographics of respondents (n=15)

| Characteristic | Sub-groups | % | Frequency |
| --- | --- | --- | --- |
| Gender | Female | 73.3 | 11 |
|  | Male | 26.7 | 4 |
| Age range | 25-34 | 26.7 | 4 |
|  | 35-44 | 33.3 | 5 |
|  | 45-54 | 13.3 | 2 |
|  | 55+ | 26.7 | 4 |
| Ethnicity | British | 26.7 | 4 |
|  | Northern Irish | 53.3 | 8 |
|  | Irish | 13.3 | 1 |
|  | Any other white British background | 6.7 | 2 |
| Years working as a GP | 0-10 years | 46.2 | 6 |
|  | 11-20 years | 23.1 | 3 |
|  | 21-30 years | 7.7 | 1 |
|  | 31+ years | 23.1 | 3 |
|  | Prefer not to say | - | 2 |
| Number of GPs within their practice | 3 | 21.4 | 3 |
|  | 4 | 28.6 | 4 |
|  | 5 | 28.6 | 4 |
|  | 6 | 14.3 | 2 |
|  | 7 | 7.1 | 1 |
|  | Prefer not to say | - | 1 |
| Location of the GP practice | Urban | 57.1 | 8 |
|  | Rural | 42.9 | 6 |
|  | Prefer not to say | - | 1 |

### GP interactions with rare disease patients

Ninety-three percent (n=13) of GPs had experience of rare disease patients in their career. When asked how often they encounter an individual with a rare disease, the responses were, 8% (n=1) ‘once a week’; 39% (n=5) ‘every month’, 31% (n=4) ‘every six months’ and 23% (n=3) said ‘every year’. Forty-six percent (n=6) of GPs surveyed said they occasionally/frequently encounter patients attending clinic with information about their rare disease / potential diagnosis / treatment options. When asked to briefly outline their interactions with rare disease patients various responses were given including, ‘*Some know lots about their condition, some not so much. I have told patients I don’t know anything about their condition in the past’* and *‘As the GP my role frequently involves being central to the co-ordination of their care’*. Sixty-seven percent (n=8) of GPs reported that they do not normally discuss genetics with rare disease patients. Sixty-nine percent (n=9) of GPs surveyed said they believe they can learn from rare disease patients. Specifically, they stated these patients provide *‘useful insights’*, have remarkable *‘coping abilities and humanity’*, unique ‘*knowledge of diseases’* and they can learn from their diagnosis journey for future patients. Fifty-four percent (n=7) of GPs believe that the expectations a rare disease patient has of them is achievable. The remaining 46% (n=6) provided various reasons as to why they felt otherwise including: lack of resources, inadequate communication from secondary care, lack of understanding and confidence.

GPs described many common frustrations among rare disease patients (Figure 1) These included: long delays, accessing services, finding a diagnosis, getting practical support, lack of follow up, lack of access to specialist services, a benefits system that cannot deal with the patients with complex conditions, distances to clinics, poor transport services, frustration, chronic pain, feeling left behind due to lack of understanding of their condition, having to repeat their history to every doctor and lack of awareness.

**Figure 1.**
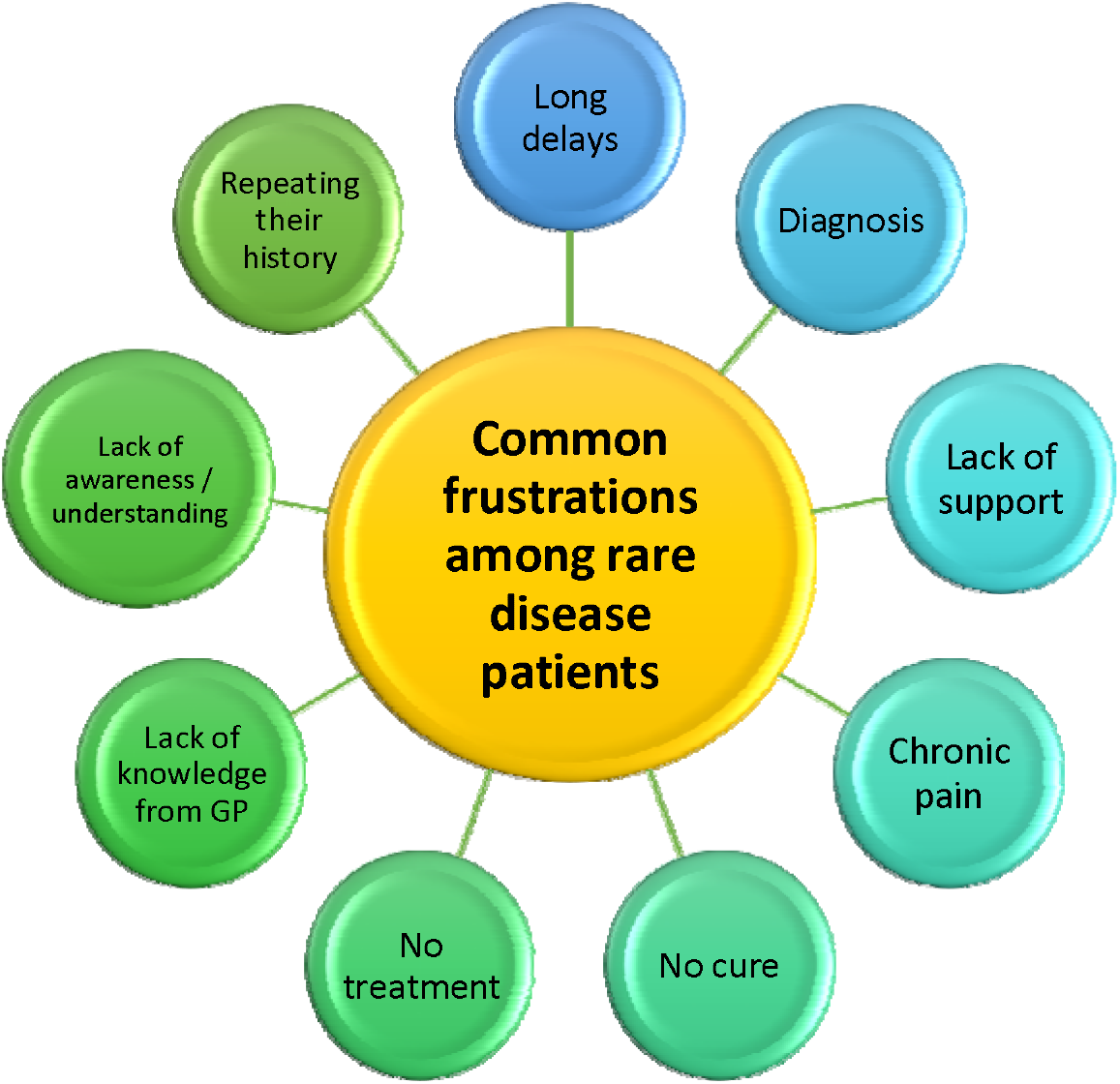
The common frustrations among rare disease patients

### Training

Seventy-one percent (n=10) of GPs said ‘rare disease’ was not included as a topic in their training to become a GP and none of them had undertaken any rare disease training since working as a GP. When asked if they felt they had been adequately trained to care for patients with a rare disease, 79% (n=11) of GPs stated ‘no’. Ninety three percent (n=13) of GPs said they would be interested in undertaking training to enable them to improve their care for rare disease patients. Several suggestions were made as to how this training should be delivered including practice-based learning sessions, workshop, online module and face-to-face training.

### Support

Fifty-seven percent (n=8) of GPs surveyed had never referred a patient to a rare disease charity. 64% (n=9) of GPs were unaware of the NIRDP, (24) the umbrella charity for rare diseases in NI. Eighty-six percent (n=12) of GPs felt there is not sufficient care/support available for those with a rare disease. When asked what more they feel could be done through charity/support groups, responses included: funding, support/specialist nurses, increased awareness and more online networking.

The word cloud (Supplementary figure 1) illustrates the biggest challenges GPs face when caring for rare disease patients. Many challenges were highlighted including: finding information, poor communication from secondary care, time constraints, lack of knowledge / resources, relying on the patient to be well informed about their condition, access to specialists and getting medication right. Despite very limited capacity, a GP stated that, ‘*Frequently there are no easy answers so the best thing is to spend time with the patient listening to concerns and offering support’*.

### Information

When GPs were asked if they encourage rare disease patients to research their condition for themselves, 85% (n=11) said they do. Thirty-nine percent (n=5) stated that patients normally bring information about their rare disease to appointments. Forty-six percent (n=6) of GPs surveyed said they do not normally give rare disease patients any literature to take away. Those that do said they get literature from the Internet, ‘recognised websites’, web mentor and Patient UK (27). ‘Recognised websites’ was not elaborated upon. One GP stated, ‘*Normally the patient knows more than I do…*’.

Figure 2 displays the top priorities that GPs suggested would help most in their interactions with patients and / or other HCPs about rare disease(s).

**Figure 2.**
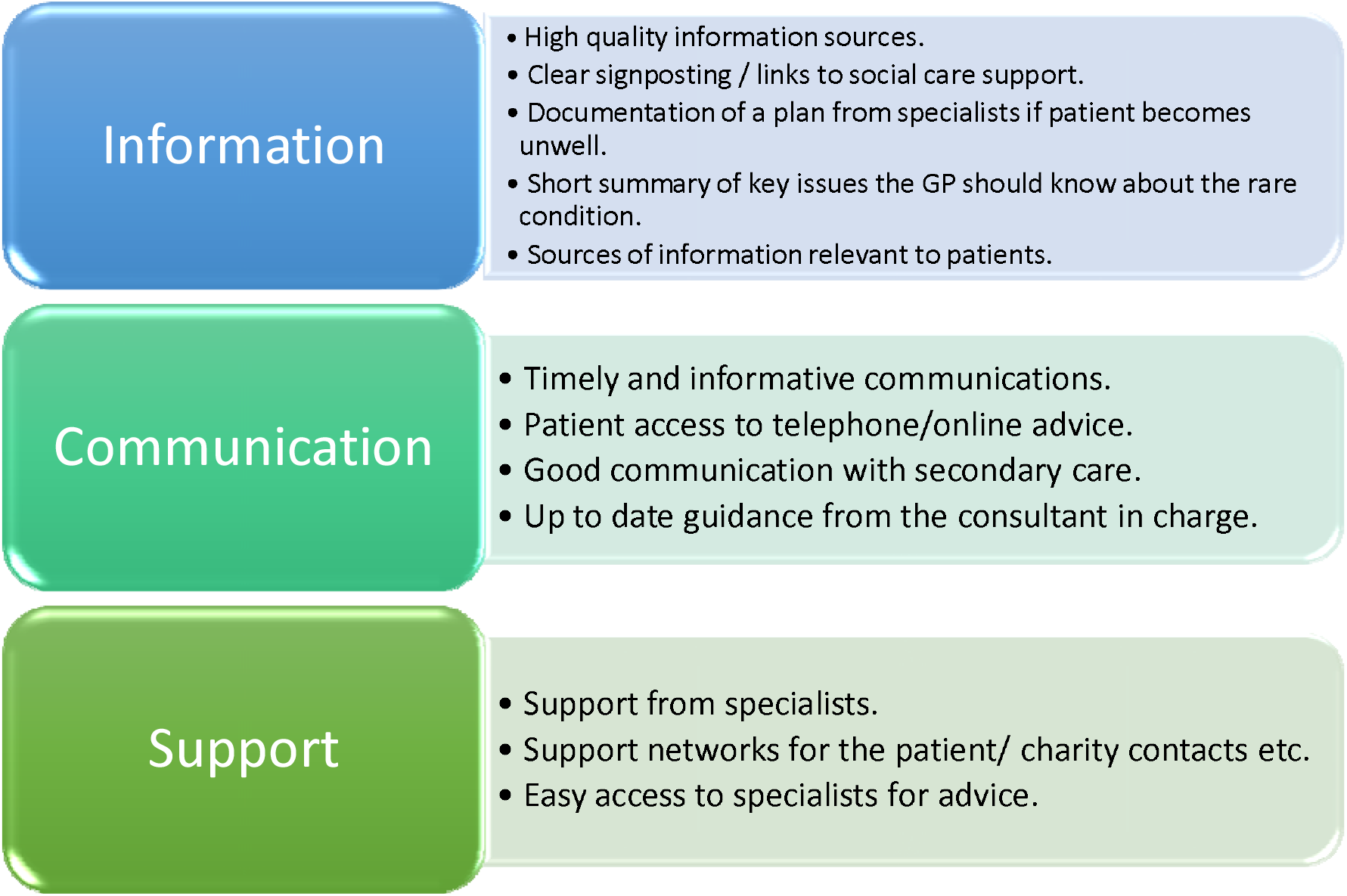
Top priorities that would help GPs most in their interactions with patients / other healthcare practitioners regarding rare disease(s).

## Discussion

### Summary

Our research identified the need for appropriate support to be developed to enable GPs to more effectively identify and care for those living with a rare disease. Training focusing on caring for rare disease patients was specifically mentioned by GPs with many never having received appropriate training as part of their vocational qualification. There was a desire for GPs to know more and to gain confidence when making decisions for this patient group’s care. For many GPs the concept of genetics is currently viewed as an element of care beyond their remit; raising awareness through training would enable them to appreciate the responsibility their specialty has within Primary Care. Currently there is no Royal College of General Practitioners e learning which focuses on rare diseases. Given the role Genomics plays in many conditions seen in primary care every day, this is a much needed area for development to link GPs to relevant national information and support sources.

An awareness around the resources for rare disease patients available online would be beneficial for GPs. Having access to such resources would enable them to provide patients with reliable information and thereby avoid endless hours of trawling the internet or the risk of lead them down the wrong path. It was also suggested that the use of databases such as Orphanet (28), OMIM (29) and Gene Reviews (30) may aid early diagnosis, thereby starting patients on the correct treatment plan at a much early stage in their journey.

Through this study it was acknowledge that GPs share a unique relationship with rare disease patients and that both parties can learn much from one another. By appreciating the value each can bring the level of care can be greatly enriched.

### Strengths and Limitations

The use of an online survey made this research more accessible for a wide audience to access remotely. The survey was promoted *via* email and Twitter; using other methods of dissemination may have helped to generate more responses. To gather more in-depth responses in the future it would be interesting to build upon this research by working with RCGP to conduct interviews with GPs to gain a greater insight into their views which is difficult to gather *via* a survey with predefined questions. In particular, it would be useful to focus future research on training for GPs. In this study, data were collected from a relatively small population group, but findings may be applicable for a wider population due to the common challenges and specific needs of GPs when caring for those with a rare disease.

### Comparison with existing literature

Previous research has indicated that a lack of knowledge among GPs causes feelings of inadequacy when caring for rare disease patients (3). It therefore seems crucial that training is considered as a priority to enable GPs to more confidently care for this unique group of patients. In agreement with the current study, GPs have previously reported their desire for additional training in this field (16, 31). Knight and Senior (2006) (9) highlighted the lack of a defined role for the GP when dealing with patients with a rare disease and their families, as well as a lack of available support and resources for GPs. Although individually rare, collectively rare diseases affect a significant proportion of the general population (32). This means, most physicians will face the diagnosis or treatment of a rare disease at some point in their professional lives (31). Given this low level of interact it is interesting that 93% of GPs surveys in our study would be interesting in undergoing training in the field of rare diseases. A study conducted last year by Hariyan *et al*. (2020) found that increased training for Healthcare Professionals *via* workshops and lectures improved their rare disease knowledge (33). Research conducted in Poland among medical students at a university reported that 95.4% of respondents perceived their knowledge about rare disease as insufficient or very poor and 92.2% did not feel prepared for caring for rare disease patients (34). Harivan *et al*. (2020) suggests it would be beneficial to commence rare disease specific training at an early stage in medical training. A lack of awareness of rare diseases among GPs has been highlighted in previous research (5, 35, 36).

A recent study by Byrne *et al*. (2020) reported that few GPs are aware of or have used Orphanet (a primary source of information on rare disease) (35). Given that the current study found that GPs turn to the Internet as a source of information for rare disease, it is imperative they are aware of reliable sources to depend on and enhance their knowledge. The Internet has been shown to play a significant role to source clinical information (15, 36) however patients have reported that GPs frequently give incorrect information (15).

Domaradzki and Walkowiak (2019) concluded that many medical students feel it is the responsibility of geneticists and paediatricians to be specifically educated and trained in the field of rare disease (34). Again, this highlights the need for early training interventions to raise awareness of the importance of understanding the needs of rare disease patients. Genomic medicine is being rolled out across all medical specialities in the UK and therefore rare diseases are no longer the domain of specialist genetic services. Smit *et al*. (2019) found that GPs believe that genomic risk may become a responsibility within primary care and they recommended a shared decision making approach to guide the testing process (37). Many adults are living with a rare disease that is not diagnosed in childhood (38).

A study in Australia conducted a survey to investigate if the healthcare needs of rare disease patients are being met. Almost three quarters of those surveyed reported having received no or not enough information at the time of diagnosis (39). This complements the current study where almost half of GPs said they do not give patients information to take away with many encouraging patients to research their condition for themselves. Similarly McMullan *et al*. (2020) found that rare disease groups expressed their desire for HCPs to interact with rare disease group’s information resources by joining consortiums, passing on relevant leaflets, as well as the usual channels (40).

Our survey found that 57% of GPs did not refer their patient to a rare disease charity and 64% were not familiar with the only national charity for rare diseases across Northern Ireland, NIRDP. Previously Anderson *et al*. (2013) discovered that 87% (n=26) of patients wished to receive information on support groups, with only 43% (n=13) receiving this (41). Creating awareness among GP communities in NI and worldwide of the charities and support groups available is key as rare disease patients find charities to provide continued support and may enable them to communicate with others who live with the same condition. The knowledge gap in rare diseases is compounded by the limited availability of information and official guidelines (42).

Our study highlighted the fact that GPs and rare disease patients share a unique relationship in which patients often take on the role as the expert. As stated previously, 69% of GPs surveyed believe they can learn from rare disease patients stating, *‘One gave me a short story written by a sufferer which was a real eye opener and very helpful’* and *‘They may have had difficulty in obtaining a diagnosis so hearing about how this came about is helpful for future pattern recognition’*. The two parties complement one another as HCPs have valuable insight into how a rare disease is exhibited, what medications do and do not work, and the true burden of the disease (43). HCPs may also identify areas for additional support, which the patient may be unaware of (44). Further research would be of value focusing solely on GPs rather than the wide group of HCPs.

The literature shows how online databases such as Orphanet may be of use to GPs in order to provide a diagnosis for the rare disease patients (2, 15). This may decrease the diagnostic delay frequently experienced and provide reliable information to GPs surrounding the diseases that they experience. Promoting awareness of the different online resources available may empower GPs to make an accurate diagnosis and provide support to their patients. One GP stated, ‘*We are on a shared journey to know more’* when talking about meeting the expectations of rare disease patients.

### Implications for research and/or practice

This study has acknowledged the difficult role GPs find themselves in when caring for patients with a rare disease. Lack of training and reliable information make providing an adequate level of care extremely challenging. Through this study the need for better training has been identified with 93% of GPs surveyed stating their desire to know more and build upon their current knowledge. Incorporating this training at an early stage in GP training appears to be the most effective and highlights the importance of being aware of this patient group and their unique needs. Awareness of rare diseases in general should be raised among the GP population with a particular emphasis on the resources available to help them as HCPs but also to benefit patients by providing them with reliable sources of information and perhaps connecting them to others living with the same conditional elsewhere in the world. The Royal College of General Practitioners in partnership with Health Education England Genomics Education Programme has developed the Genomics Toolkit (45). This is used to support increasing understanding, raise awareness of Genomics Medicine and support primary care with increased knowledge of how genomics can contribute to improving patient care in a ‘genomics era’. Raising awareness of this resource would be helpful alongside developing national specific resources. Furthermore, the use of online databases for rare disease patients enhances the care provided and can help to reduce the diagnostic odyssey that many patients find themselves living in.

## Conclusion

New insights have been added to the area of rare disease care and the need for training, support and more awareness have been highlighted. As echoed in previous research within our research team, this study recognises the importance of listening to all voices within the rare disease community (40, 46). The surveys have provided insight into the interactions GPs have with rare disease patients in NI and have identified where more support is needed to enable GPs to carry out this challenging role.

There appears to be a gap in the information and training available to GPs and this issue needs to be combatted to satisfy the aims of the *Northern Ireland Rare Disease Implementation Plan (2015)* (17) and therefore a clear area to target would be accessible training. GPs require more effective and convenient training programs and a universal web-based resource would be favourable among the GP communities in NI who are searching for information online.

Easing communication between GPs and hospital specialists is of paramount importance to give GPs access to the best information on how to treat their rare disease patients. Awareness should be generated to existing charities and support groups for rare disease patients as this eases the experience for the patient and removes pressure from the GP. A quick reference guide for rare disease was recently developed to help support rare disease management in general practice (15). A better infrastructure should be introduced in order to allow smooth communication and to keep records of up to date information and care plans accessible to all involved in the patients care. It is evident that GPs do their best to care for rare disease patients in what are often very difficult circumstances with no hope of effective treatment or cure, minimal (if any) local specialists to rely on, and sub-optimal training to equip them with tailored ways to effectively manage each rare disease.

## Data Availability

The datasets generated and/or analysed during the current study are not
publicly available due as they contain potentially identifable information but
are available from the corresponding author on reasonable request.

## Ethical approval

Ethical approval was provided by the Faculty of Medicine, Life and Health Sciences, Queen’s University of Belfast (QUB) research ethics committee (FMHLS 18.47v3).

## Acknowledgements

We thank all GPs who participated in this study. We also thank NIRDP (www.nirdp.org.uk) for helping design the survey and promote the study.

## Supplementary file

*Supplementary file 1*

- Template survey
- PDF

*Supplementary figure 1*

- The biggest challenges GPs face when caring for rare disease patients.
- PDF

