## Supplementary file for "Perceptions and experiences of rare diseases among General Practitioners: an exploratory study"

### File S1

#### GP's perception and experience of rare diseases

#### Survey

**Introduction**

The aim of this project is to gain an understanding of GP's perception and experience of rare diseases. We will also work with the Northern Ireland Rare Disease Partnership to explore service user (individuals living with a rare disease(s)) perspectives of the GP-patient partnerships. It is hoped that increasing awareness around this issue will in turn improve the care provided for those living with a rare condition.

This survey should take about 15 minutes to complete. Results from this survey will be collated anonymously and used to increase awareness around this issue will in turn improve the care provided for those living with a rare condition.

It is not essential to answer every question! Complete survey data is most useful to us, but we understand you may not wish to disclose (or have time to complete) some information – please feel free to skip over any such question(s).

The survey is available in other formats (large print, hard copy, verbal delivery, and different colour contrasts). Please contact if you wish to access these.

**Many thanks for your participation; it is much appreciated!**

**By completing this survey, I am confirming that I understand the purpose of the study and give my consent for my responses to be used anonymously for research.** (*Please tick*)

**Yes** □ **No** □

##### Section A - General information

*This information is gathered so we can evaluate whether our responses are from a diverse range of individuals. We are keen to maximise inclusion! (please tick)*

1. What is your gender? 15 answered

| |
|---|
| Male |
| Female |
| Prefer not to say |
| Prefer to describe myself as: |

2. What age bracket best describes you? 15 answered

| |
|---|
| 25-34 |
| 35-44 |
| 45-54 |
| 55+ |
| Prefer not to say |

3. What is your ethnic origin? 15 answered

| **White British** | **Asian or Asian British** |
|---|---|
| British<br>English<br>Northern Irish<br>Scottish<br>Welsh<br>Any other white British background? If so, what? ___________________________ | British Asian<br>Indian/British Indian<br>White & Asian<br>Bangladeshi/British Bangladeshi<br>Any other Asian background, if so, what?<br>________________________________________ |
| **Other White European** | **Black or Black British** |
| Irish<br>Cypriot<br>Greek<br>Greek Cypriot<br>Turkish<br>Turkish Cypriot<br>Italian<br>Polish<br>Kosovan<br>Albanian<br>Bosnian<br>Kurdish<br>Any other White Background, if so what?<br>_____________________________________ | Black British<br>African<br>Caribbean<br>Nigerian<br>Somali<br>Any other black origin, if so, what?<br>_________________________________________ |
| **Mixed** | **Other Ethnic Groups** |
| White & Black Caribbean<br>White & Black African<br>White & Asian<br>Any other mixed background, if so what?<br>_____________________________________ | Chinese<br>Jewish<br>Iranian<br>Arab*<br>Latin American*<br>North African*<br>*Specify: ____________________ |

4. May we contact you about a potential interview to follow up on responses in this online survey? *(described above)* 15 answered
   Yes □ No □

If yes, please provide your email address (preferred contact details, ideally email address?) below:

______________________________________________________________________________

**Section B – Survey**

*This section of the survey is divided into three sections: Interactions with rare disease patients, Contact with charity/support groups and Training (please tick or provide further details when prompted).*

***Interactions with rare disease patients***

5. How many years have you been working as a GP? 13 answered, 2 preferred not to say

| |
|---|
| 0-10 years |
| 11-20 years |
| 21-30 years |
| 31 years or more |

6. How many GPs work in the practice where you are currently based? 14 answered, 1 preferred not to say

______________________________________________________________

7. Is the practice you work in based in an urban or rural location? 14 answered, 1 preferred not to say

| |
|---|
| Urban |
| Rural |

8. Have you had any experience of rare disease patients so far in your career? (If no please continue to question 16) [use smart survey to skip questions depending on response given] 14 answered, 1 preferred not to say

| |
|---|
| Yes |
| No |

9. Approximately how often do you encounter an individual with a rare disease? 13 answered, 2 preferred not to say

| |
|---|
| Once a week |
| Every month |
| Every six months |
| Every year |

10. And following from question 9: Do patients normally bring information about their rare disease to an appointment? How often do you encounter patients attending clinic with information about their rare disease / potential diagnosis / treatment options? 13 answered, 2 preferred not to say

| |
|---|
| Never |
| Rarely |
| Occasionally |
| Frequently |
| N/A |

11. Please outline briefly your interactions to date with rare disease patients (including details of what rare diseases you have encounters, age of patients, the relationship between yourself and those patients, the knowledge the patient had of their disease) 12 answered, 3 preferred not to say

______________________________________________________________________________

12. What is the biggest challenge you face when dealing with a patient with a rare disease? 12 answered, 3 preferred not to say

13. What frustrations are common among rare disease patients? 12 answered, 3 preferred not to say

14. Do you normally discuss genetics with rare disease patients? 12 answered, 3 preferred not to say

| |
|---|
| Yes |
| No |

15. Do you encourage patients to research their condition for themselves? 13 answered, 2 preferred not to say

| |
|---|
| Yes |
| No |

16. Do you normally give the rare disease patient any literature to take away? 13 answered, 2 preferred not to say

| |
|---|
| Yes |
| No |

If yes, where do you normally get this literature from?

17. Do you feel the expectations a rare disease patient has of you are achievable? 13 answered, 2 preferred not to say

| |
|---|
| Yes |
| No |

If no, why not?

18. Do you feel you can learn from rare disease patients? 13 answered, 2 preferred not to say

| |
|---|
| Yes |
| No |

If yes, in what way?

____________________________________________________________________________
____________________________________________________________________________

19. What are the top (up to three) priorities that would help you most in your interactions with patients/other healthcare practitioners about rare disease(s)? 12 answered, 3 preferred not to say

1._____________________________________________________________________
2._____________________________________________________________________
3._________________________________________________________________

**Contact with charities/support groups**

20. Have you ever referred a patient to a rare disease charity? 14 answered, 1 preferred not to say

| |
|---|
| Yes |
| No |

If yes, which charity have you used most often?

____________________________________________________________________________
____________________________________________________________________________

21. Are you aware of the Northern Ireland Rare Disease Partnership? 14 answered, 1 preferred not to say

| |
|---|
| Yes |
| No |

If yes, have you ever contacted them?

| |
|---|
| Yes |
| No |

22. Do you think sufficient care/support is available for those with a rare disease? 14 answered, 1 preferred not to say

| |
|---|
| Yes |
| No |

If no, what more do you think could be done through charity/support groups?

____________________________________________________________________________
____________________________________________________________________________

**Training**

23. Was ‘rare disease’ included as a topic in your training to become a GP? 14 answered, 1 preferred not to say

| |
|---|
| Yes |
| No |

24. Have you undertaken any rare disease training since working as a GP? 14 answered, 1 preferred not to say

| |
|---|
| Yes |

| |
|---|
| No |

If yes, please outline briefly what this training involved and how it was delivered.

________________________________________________

________________________________________________

25. Do you feel you have been adequately trained to deal with patients with a rare disease? 14 answered, 1 preferred not to say

| |
|---|
| Yes |
| No |

26. Would you be interested in undertaking training to enable you to deal with rare disease patients in a more efficient way? 14 answered, 1 preferred not to say

| |
|---|
| Yes |
| No |

If yes, what way would you prefer to receive this training, i.e. online module supported by RCGP, workshop etc.?

________________________________________________

________________________________________________

**Thank you for taking the time to complete this survey. We very much appreciate your input - you are helping to improve services for individuals affected by rare diseases in Northern Ireland.**

**If you wish to provide further details/or have any questions about this survey, please contact using 'Survey GPs' in the subject line.**

We are also keen to interview a subset of people who complete this survey to obtain further information; this interview may take 15 minutes – two hours depending on how much you wish to say. If you are happy to be contacted, please do supply your email address in question 4. We appreciate and welcome your input to this project. If you wish to provide further details and / or have any questions about this survey, please contact using 'Survey GPs' in the subject line.

**Thank you, this survey is now complete**

### Figure S1

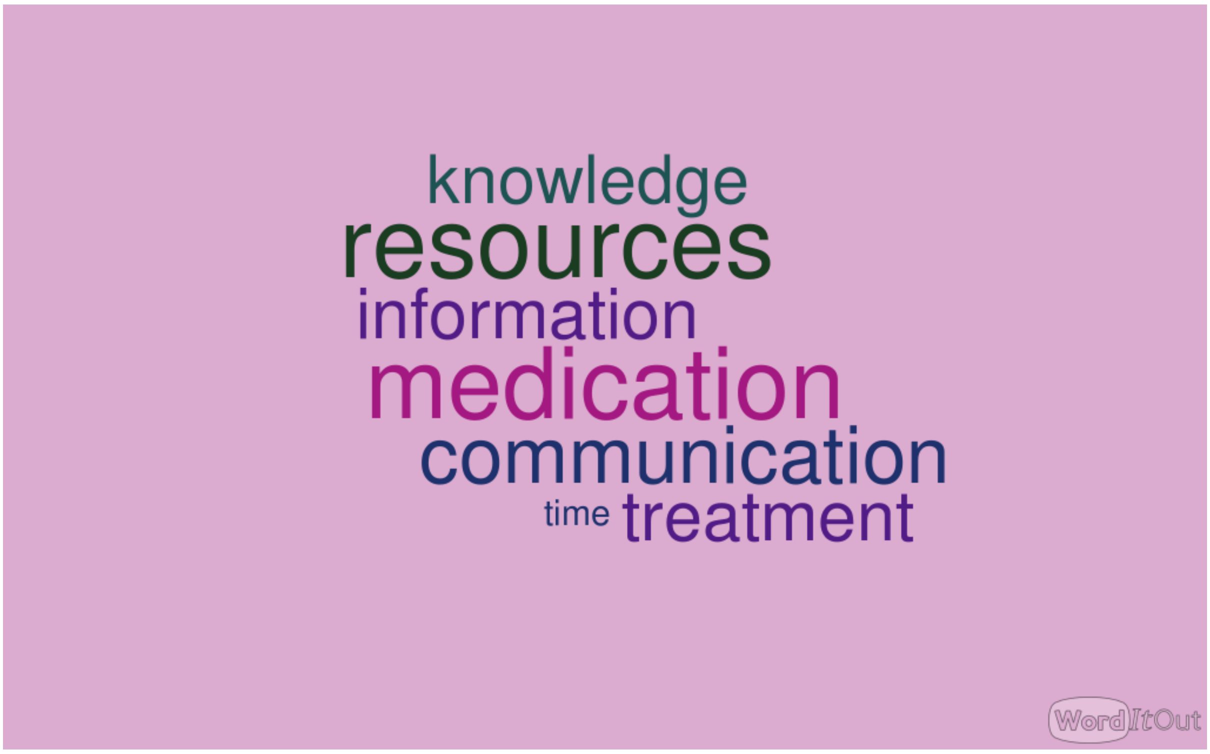

**Figure S1**

The biggest challenges GPs face when caring for rare disease patients.
